# Machine learning predicts treatment response to nusinersen in non-sitter Spinal Muscular Atrophy (SMA)

**DOI:** 10.1101/2025.10.22.25337435

**Authors:** Georgia Stimpson, Emer O’Reilly, Giorgia Coratti, Deborah Ridout, Tapabrata Chakraborti, Robin Mitra, Roberto de Sanctis, Marika Pane, Mariacristina Scoto, Eugenio Mercuri, Francesco Muntoni, Giovanni Baranello, the International SMA Consortium

**Affiliations:** Dubowitz Neuromuscular Centre, UCL Great Ormond Street Institute of Child Health, London, UK; Health and Medical Science Programme, The Alan Turing Institute, London, UK; NIHR Great Ormond Street Hospital Biomedical Research Centre, UCL Great Ormond Street Institute of Child Health, London, UK; Paediatric Neurology Unit, Catholic University, Rome, Italy; Centro Clinico Nemo, U.O.C. Neuropsichiatria Infantile Fondazione Policlinico Universitario Agostino Gemelli IRCCS, Rome, Italy; Population, Policy & Practice Department, UCL Great Ormond Street Institute of Child Health, London, UK; UCL Cancer Institute and Dept of Medical Physics & Biomedical Engineering, UCL, London, UK; Department of Statistical Science, UCL

## Abstract

1.1

**Background:** Nusinersen has substantially increased survival and improved disease progression in Spinal Muscular Atrophy (SMA) patients. However, treatment response is heterogeneous, with some patients gaining the ability to sit and walk whilst others display less obvious changes.

**Method:** From the SMA Reach UK and the Italian Telethon Network, 124 non-sitter SMA patients treated with nusinersen under 4 years were included and randomly allocated to training and testing (80/20%). Tree and regression-based machine learning for survival outcomes were compared, and oblique random forests were selected, with a testing C-Index of 0.74. The features were selected from items of the CHOP-INTEND and HINE-2 motor function assessments, respiratory and swallowing status using mutual information.

**Findings:** Sixty-two patients (50%) achieved sitting, at a median age of 2.4 years. The predicted median time-to-sitting in those requiring tube feeding at treatment initiation was 4-months later than those without, and this was the most influential factor. Specific motor function features, including the ability to kick in supine, were strongly associated with a higher likelihood of sitting.

**Interpretation:** This work provides a framework for predicting nusinersen-response and represents the first stage in personalised counselling on treatment plans for SMA.

**Funding:** Support for SMA Reach is provided by Biogen, Roche and Novartis, and historically NIHR BRC, SMA Trust, the MRC Translational Research Centre and MD UK. Support for the Italian network is provided by Famiglie SMA, Telethon (GSP 13002), and ASAMSI.

**Research Into Context:** *Evidence before this study:* A literature search on PubMed up to January 1^st^ 2025 with the term “(“Spinal Muscular Atrophy” OR “SMA”) AND (“Nusinersen” OR “Spinraza” OR “ISIS-SMNRx” OR “ISIS 396443”) AND (“predict*”)” across all fields was performed. Achievement of sitting after nusinersen treatment in SMA 1 has been linked to the age of treatment, symptom onset and general motor function severity (CHOP-INTEND and HINE-2 score), but no multivariable analysis or prediction of outcomes after treatment was found.

*Added value of this study:* Our study identifies key patient features that are linked to nusinersen treatment response in non-sitter SMA 1 patients. This work provides a higher level of granularity beyond previous univariate models, yielding a framework for predicting personalised patient sitting. Crucially, we highlight that bulbar impairment as a proxy for disease severity is very influential in predicting a patient’s likelihood of sitting, along with specific motor function abilities at baseline.

*Implications of all the available evidence:* Despite considerable evidence of efficacy, this work provides insights into the homogenous gross motor function response observed in SMA patients treated with nusinersen, which has been under researched. The results of this study provide individualised predictions of the likelihood of patient’s outcomes after treatment with nusinersen. Crucially, this work can inform clinicians in their discussions on treatment-response with parents and carers of babies with SMA.

## 2 Introduction

Spinal Muscular Atrophy (SMA) is an autosomal recessive disease with an incidence between 1 in 6,000 and 1 in 20,000 live births ^1^. It is caused by mutations in the survival motor neuron (*SMN*) 1 gene, leading to low levels of the SMN protein. The paralog *SMN2* gene produces 10% functional SMN protein. SMA I is the most common SMA phenotype, with 60% of newborns with paediatric onset having SMA I ^2^. Patients with SMA I typically have onset of symptoms before 6 months ^3^, display a monotonic and global motor function decline with age ^4^ and if untreated never achieve independent sitting. Patients with SMA I also experience bulbar dysfunction ^5^, disordered growth ^6^ and respiratory failure ^7^.

The disease trajectory of SMA patients, and in particular of those with SMA I has been significantly altered by the advent of three disease-modifying therapies (DMT): nusinersen, onasemnogene abeparvovec (OA) and risdiplam. Nusinersen is an intrathecally injected antisense oligonucleotide which affects the *SMN2* gene to produce functional SMN protein. In the nusinersen clinical trial, ENDEAR (clinicalTrials.gov: NCT02193074) there was a hazard ratio of 0.37 for the survival in nusinersen-treated as opposed to the treatment-naive patients ^8^. There is significant variability in SMA I patient response to nusinersen, with some patients having no or limited functional gain at 6 months whilst others show variable achievement of motor milestones like head control, rolling, sitting and standing ^9^. The use of nusinersen in pre-symptomatic patients in the NURTURE trial (clinicalTrials.gov: NCT02386553) has demonstrated that acquisition of independent walking is also possible if treatment is initiated sufficiently early ^10^.

Achievement of sitting is correlated with age at treatment initiation, with-all presymptomatic patients in the NURTURE study achieving the ability to sit at a median age of 7.9 (95% CI: 5.9–9.2) months ^11^, which is within the WHO age-appropriate time window^12^. Shorter disease duration (i.e. the time between symptom onset and treatment initiation) has also been linked to improved nusinersen efficacy ^13^. Significantly higher median HINE-2 and CHOP-INTEND scores (2 vs 1, p<0.01 and 35.5 vs. 26.5, p<0.05, respectively) at baseline were observed in those who had achieved sitting within 14 months of treatment ^14^. This same analysis demonstrated that those with a HINE-2 of ≥2 at baseline had a 300% increased likelihood of sitting compared to those with a HINE-2 of 1 or 0 ^14^.

Machine learning has two key advantages compared to traditional statistical methods (i.e. parametric regression) that can be helpful in providing a prediction model of outcome in SMA patients treated with DMT. Firstly, many machine learning algorithms contain a degree of regularisation or feature selection, which allows the reduction of a high-dimensional feature space. Additionally, tree-based methods are of particular interest as they can capture non-linear relationships between features and outcomes (in this case the time to independent sitting).

### 2.1 Aims

The aim of this study was to generate a predictive model for the likelihood of sitting in non-sitter SMA patients treated with nusinersen, which was the first approved DMT for SMA. A secondary aim was to describe how this model generates predictions, with a particular interest in the interaction between individual characteristics, which have not been adjusted for in previous research.

## 3 Methods

The data utilised in this observational study was collected by the SMA REACH-UK (clinicaltrials.gov ID: NCT03520179) and Italian SMA networks, which form part of the International SMA Consortium (iSMAc)^15^. The study was approved by the SMA REACH-UK steering committee. Data was collected both in the UK and Italy using the standardised iSMAC forms.

### 3.1 Participants

Non-sitter SMA participants who had received nusinersen as their initial treatment in the UK and Italy in an iSMAc centre before December 2022 (the cut-off for data collection) were considered for this analysis. The visit closest to treatment initiation, and between 6 months before and three months after treatment, was considered as the baseline. This is consistent with the encoding that has been used for the managed access agreement of nusinersen in the UK. Sitting ability was assessed using the WHO definition of sitting ^16^ (90% of assessments), or in assessments where this was not available, a score of at least a 3 or 4 on item 5 of the HINE-2 or a score of 2 on item 1 of either the RHS or HFMSE was considered. When sitting ability was not reported at all after treatment, participants were removed. The data-cleaning process is described in Figure 1.

**Figure 1.**
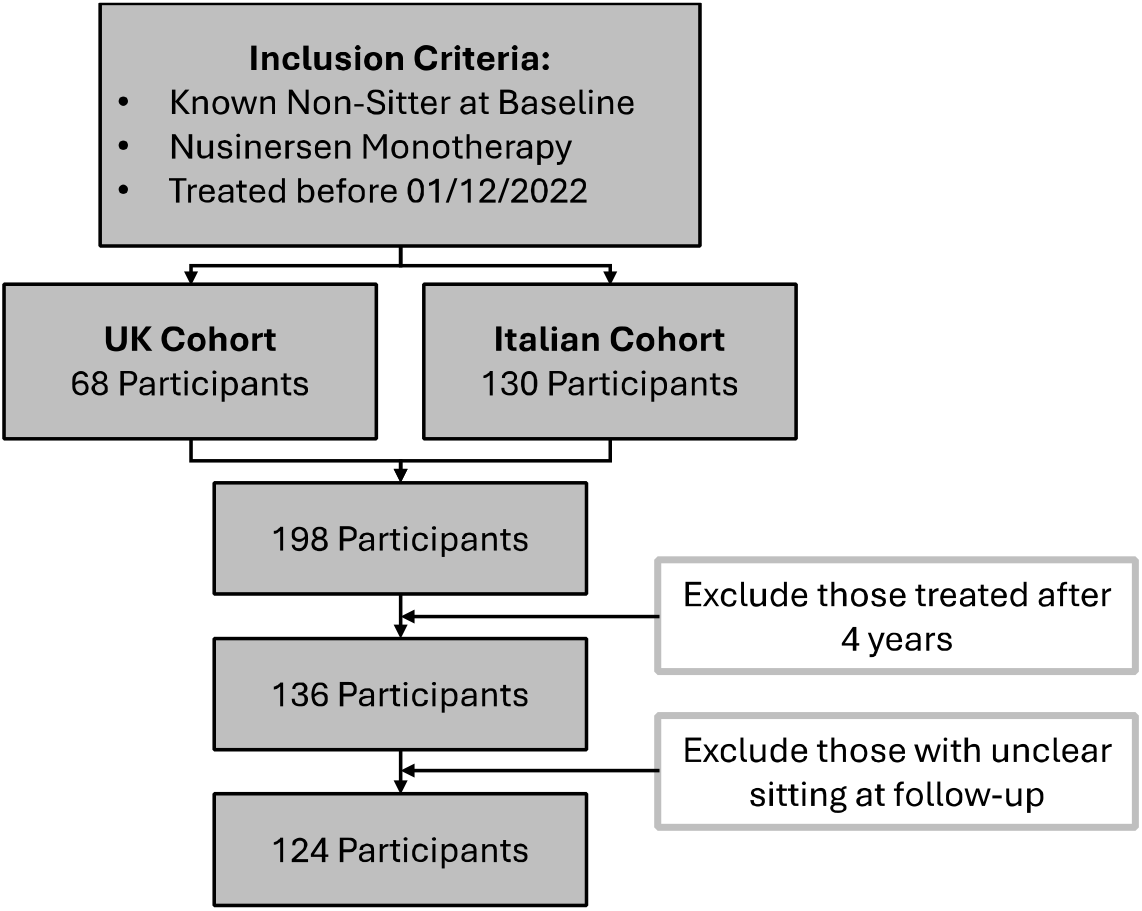
Data Cleaning Process.

### 2.2 Features

Participant gender, SMN2 copy number, nusinersen start age, and symptom onset were considered for inclusion in the model. Bulbar involvement was captured through the presence of swallowing problems and feeding support at baseline (a percutaneous endoscopic gastrostomy (PEG) surgery, gastrojejunostomy tube insertion surgery, gastrostomy tube insertion or a nasogastric tube insertion). Abnormal growth was captured through participant height, weight and BMI centile for age. Respiratory involvement was captured through an indicator for tracheostomy surgery, salbutamol initiation and NIV use (BIPAP, CPAP, “other” NIV and prophylactic NIV, but not those who used NIV only when sick) and NIV hours.

The Children’s Hospital of Philadelphia Infant Test of Neuromuscular Disorders (CHOP-INTEND) is a validated scale for patients with SMA ^17^. The scale contains 16 items, with most items scored 0-4, with a total score of 64. In some items, it is not possible to score a 1 and/or a 3. The Hammersmith Infant Neurological Examination part 2 (HINE-2) was initially developed to assess neurological motor in preterm and newborn infants ^18^, and contains 8 items, scored 0-3, 0-4 or 0-5 leading to a total score of 26. The items relate to a range of motor function milestones: head control, sitting, voluntary grasp, kicking (in supine) rolling, crawling/bottom shuffling, standing and walking. For each of these scores, the ordinal item scores were discretised into binary scores for each level (i.e. a feature for ability to score at least a 1 on a specific item). Here, if less than 5 participants scored on a single subscore, this feature was removed.

### 2.3 Outcome

The outcome of interest was time to independent sitting from first nusinersen dose. As this outcome is predominantly recorded as a longitudinal binary variable, time to sitting was interval censored between the last non-sitting and first sitting assessment time. Participants who initiated a secondary therapy (zolgensma or risdiplam) were censored at 4 months after last nusinersen dose (in line with correct scheduling), or at the first non-nusinersen dosing, whichever was earlier. Age-appropriate sitting was assessed as sitting before 9.2 months, as defined by the WHO Motor Development Study ^19^.

### 2.4 Statistics

There was a significant degree of missing data; 42% of data for height centile was missing, whilst HINE-2 was missing in 24% and feeding support in 15%. Additionally, patterns of structured missingness in line with a missing completely at random-weakly structured (MCAR-WS ^20^) and missing at random-weakly structured (MAR-WS ^20^) pattern were observed in the medical and physiotherapy fields respectively. As such, 25-fold Multivariate Imputation by Chained Equations (MICE) imputation was performed, using the method which induced the lowest error when structured missingness was simulated in the complete cases across four domains. This is described in depth in Supplemental Figure 1.

The MLR3 universe of packages in R version 4.2.2 was used to build the machine learning pipelines. The data was divided into a training sample and a holdout sample, with 20% of data randomly allocated for testing. This yielded 28 patients, and as such the final sample size was 96 patients.

Due to the high number of features relative to the number of participants, five filtering methods were considered: correlation, correlation-adjusted regression survival (CARS), mutual information maximisation (MIM), minimum redundancy maximal relevancy (MRMR) and double input symmetrical relevance (DISR). The complete feature ranking by filtering method is available in Supplemental Table 2.

Tree-based (decision trees, decision random forests and oblique random forests), causal inference trees-based (trees and random forests) and regularised regression-based (lasso, ridge and elasticnet) models for survival outcomes were considered. We utilised every measure of node impurity that had been implemented for each tree-based method, resulting in 16 total models. This is detailed in Supplemental Table 3.

Hyperparameter tuning was performed across five domains: resampling of splits, splitting criteria, stopping criteria, the number of candidate features and the number of trees, with the later two available only in the random forest algorithms. Random search with 25 hyperparameter sets generated from the parameter space was utilised. The performance of each hyperparameter set was evaluated with a 5-fold cross-validation loop. This is identical to the procedure described in Spooner et al.,2020 ^21^. Details of the hyperparameters selected for each model, and their tuned domain is available in Supplementary Table 4.

Model performance was assessed using Harrell’s C-Index, a concordance-based approach to assessing survival model goodness-of-fit. A grid search across all 16 models was performed, across all 80 features and the best 40 and 20 features for each filtering method. The C-index was calculated using 5-fold cross validation, and the two methods with the highest average C-index were pursued for a closer grid search. This was the Accelerated Oblique Random Survival Forest (AORSF) model and the ranger model with 20 features. A grid search of width five was then used to identify the neighbourhood with the highest average C-index, and in this region a grid search of width 1 was performed. This process is described fully in Figure 2, and model performance is summarised in Supplemental Figure 2 and Supplemental Figure 3. As both AORSF and Ranger (with Logrank) are tree-based algorithms, the integrated Brier score (IBS) was also considered, and the model that minimised the IBS was selected. The best method when assessed by mean IBS was the AORSF using the MRMR method with 18 features (mean IBS=0.135 (SD= 0.018), mean C-index 0.783 (SD=0.022)). The out-of-bag C-index was on average 0.742 (SD=0.086) for the selected method.

**Figure 2.**
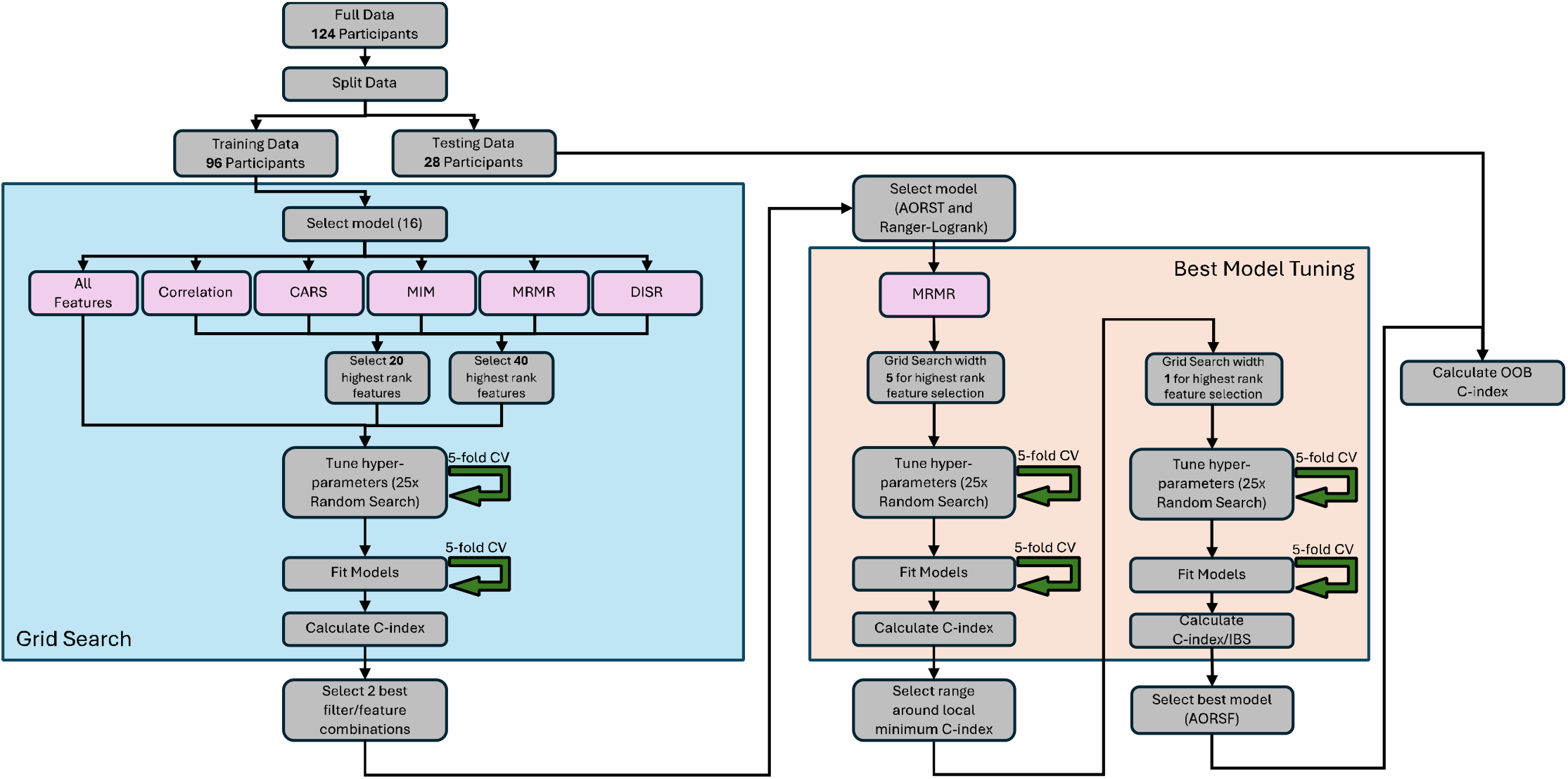
Workflow of machine learning process for the model

Variable importance was calculated using the permutation method ^22^. Here, for each feature, the difference in a given metric between the model fitted on the full data, and the model fitted on the data with that feature randomly permuted is calculated. This is calculated in out-of-bag samples. Partial dependence metrics were used to summarise the population-level impact of features on the overall model predictions ^23^. On the patient specific level, the survival-specific explainability outcome SurvSHAP was utilised, which provides insights into how the addition of individual features effects the survival risk over time ^24^. Notably, SurvSHAP values were developed with traditional survival analysis in mind, in that a positive value indicates an increased survival time and a lower probability of death. However, in this analysis the interpretation is flipped-increased time-to-sitting is associated with a lower probability of sitting and a negative outcome. Therefore, a negative SurvSHAP value is associated with an earlier predicted time-to-sit. Local Interpretable Model-agnostic Explanations (LIME) is an explainability outcome that explains the prediction neighbourhood, and the SurvLIME is a survival-specific implementation of the LIME process ^25^.

## 4 Results

The inclusion criteria yielded a total sample size of 124 participants, who had a median of 7 assessments each, for an average follow-up time of 2.6 (IQR: 1.5, 4.1) years. Participant demographics are summarised in Table 1.

**Table 1.** Summary Table of Baseline Characteristics by Training and Testing Dataset.

| Med (IQR) |  | Full Cohort | Training Cohort | Testing Cohort |
| --- | --- | --- | --- | --- |
| N |  | 124 | 96 | 28 |
| Symptom Onset Age (months) |  | 4.2 (2, 6.4) | 4.7 (2, 6.6) | 3.7 (2.3, 5.3) |
| Nusinersen Start Age |  | 6.8 (3.8, 11.5) | 7.4 (4, 12.4) | 6 (3.2, 10.3) |
| Sex | F | 53 | 37 | 16 |
|  | M | 71 | 59 | 12 |
| SMA Type | 0 | 1 | 1 | 0 |
|  | I | 110 | 82 | 28 |
|  | II | 6 | 6 | 0 |
|  | Pre-symptomatic | 7 | 7 | 0 |
| SMN2 Copy Number | 1 | 3 | 2 | 1 |
|  | 2 | 92 | 69 | 23 |
|  | 3 | 21 | 19 | 2 |
|  | 4 | 1 | 1 | 0 |
| CHOP-INTEND Total Score |  | 29 (22, 39) | 30 (23, 39) | 25 (20, 37) |
| HINE-2 Total Score |  | 2 (1, 3) | 2 (1, 4) | 2 (1, 3) |
| Feeding Support | No | 62 | 48 | 14 |
|  | Yes | 43 | 33 | 10 |
| Swallowing Difficulties | No | 69 | 55 | 14 |
|  | Yes | 27 | 19 | 8 |
| Respiratory Support | None | 51 | 40 | 11 |
|  | NIV | 43 | 34 | 9 |
|  | Tracheostomy | 13 | 10 | 3 |
|  | NIV Hours | 10 (8, 11) | 8 (8, 10) | 10 (10, 14) |
| Weight Centile |  | -1.61 (-2.78, -0.36) | -1.69 (-2.93, -0.62) | -1.4 (-2.04, 0.45) |
| Height Centile |  | -0.76 (-2.40, 0.51) | -0.92 (-2.41, 1.03) | -0.75 (-2.1, -0.37) |
| BMI Centile |  | -1.42 (-2.57, -0.43) | -1.75 (-2.93, -0.62) | -1.31 (-2.03, 0.12) |

In total, 62 (50%) of the cohort achieved independent sitting, and the median age of sitting using the Kaplan-Meier estimate was 2.41 years (95% CI: 1.8, NA). Age-appropriate sitting acquisition was observed in only 5 (4%) participants, and this consisted of 2 participants treated pre-symptomatically (before 3 months, both with baseline CHOP-INTEND of 60), and 3 SMA type Ic. The complete Kaplan-Meier curve is shown in Figure 3.

**Figure 3.**
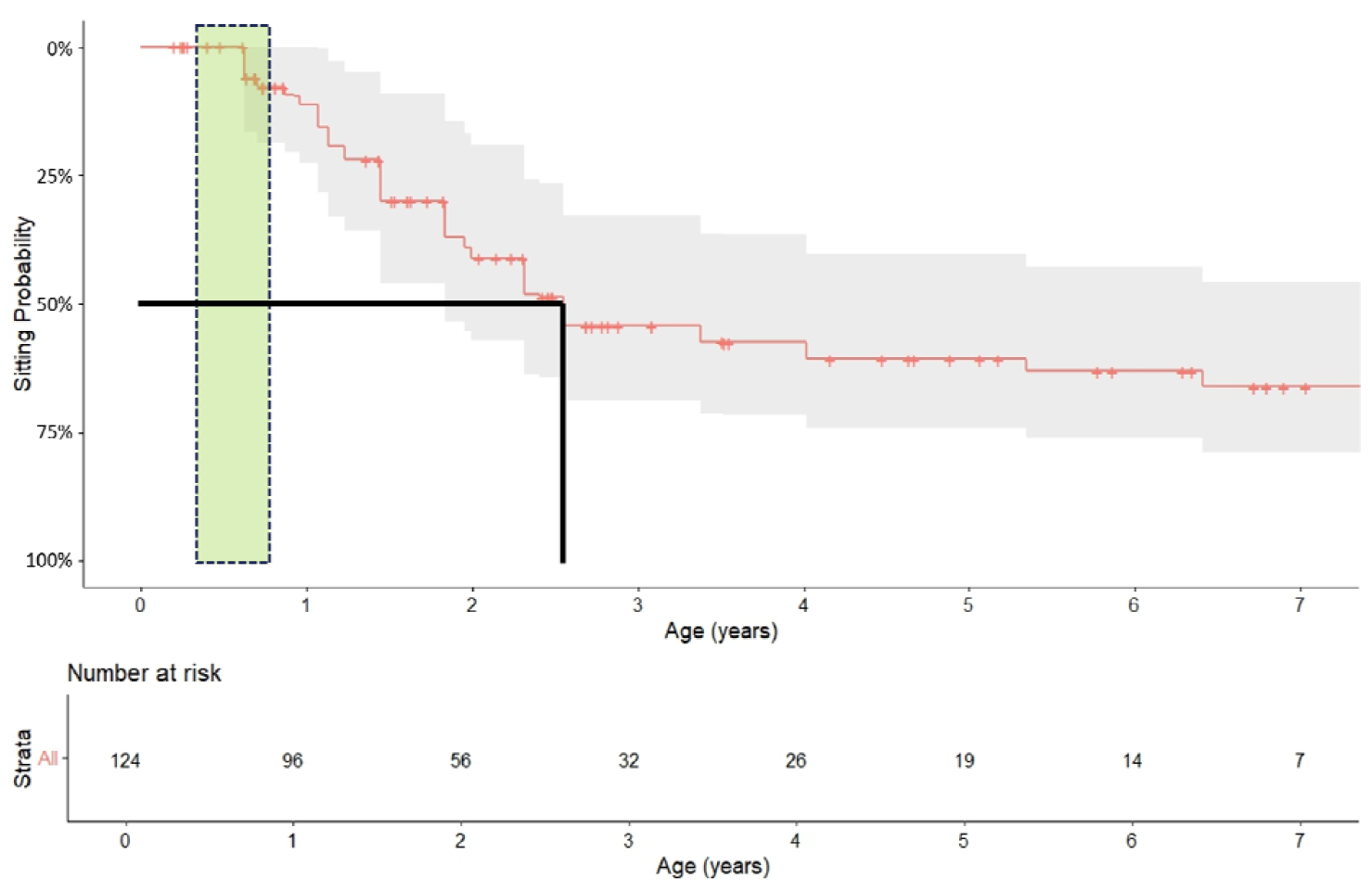
Age at Sitting in Cohort with 95% CI, with vertical dashed green box indicating the age range associated with age-appropriate sitting, and the black continuous line demonstrating the age at median sitting.

Overall model selection was performed by optimising the 5-fold cross validated Harrel’s C-index and integrated Brier score on the training data across model type and filtering selection method. The model which gave the best training performance was the Accelerated Oblique Random Survival Forests (AORSF) model with the minimum redundancy maximal relevancy (MRMR) method of feature selection with 18 features.

The CHOP-INTEND total score and HINE-2 total score were selected for inclusion in the model, along with the ability to kick as scored on the HINE-2 (0 vs 1/2/3/4). In terms of CHOP-INTEND items selected to be included in the model, these were as follows: antigravity shoulder movement where the shoulder is lifted (0/1/2/3 vs 4 on item 1), any lower limb movement proximal to the ankles (0/1 vs 2/3/4 on item 2), maintenance of hand grip whilst traction is applied and shoulder lifted (0/1/2/3 vs 4 on item 3), rolling fully prone from supine (0/1/2 vs 3/4 on item 7), antigravity arm movement whilst in side lying (0/1/2/3 vs 4 on item 8) and activation of the neck muscles during a pull to sit motion (0 vs 2/4 on item 14). Diagrams which describe these specific motor function abilities are available as Supplementary Document 1. Beyond motor function, nusinersen start age, symptom onset age, feeding assistance, swallowing problems, tracheostomy ventilation, growth centiles (BMI, height and weight) and NIV hours were selected for inclusion in the model. As assessed by permutation importance using the C-index, the 7 most important features in the model were the indicators for feeding support and swallowing difficulties, BMI, antigravity shoulder movement where the shoulder is lifted, muscle activation of the neck muscles during a pull to sit motion and rolling fully prone from supine. The full importance of all features as assessed by permutation is shown in Figure 4.

**Figure 4.**
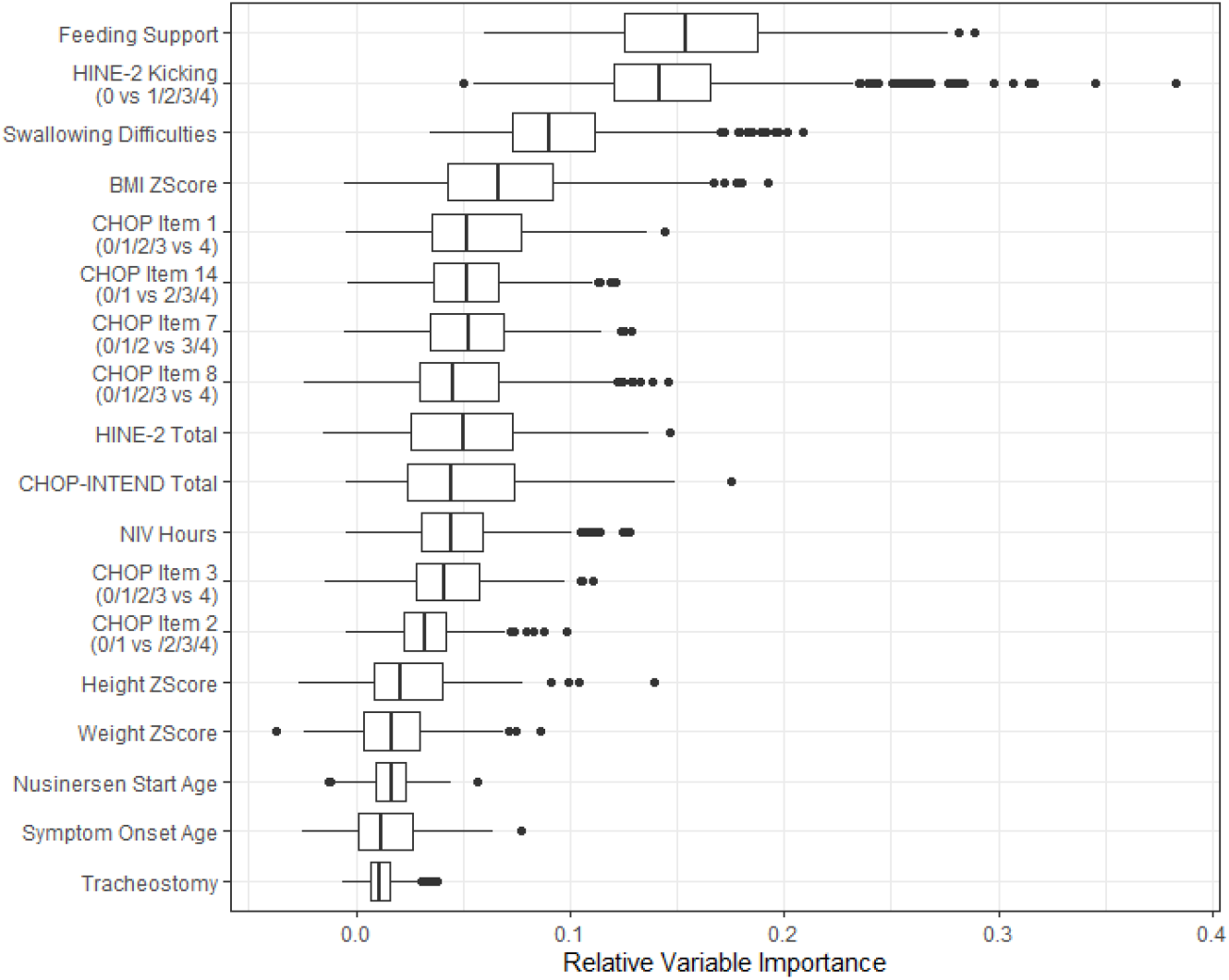
Feature Variable Importance by Permutation

The presence of feeding support and swallowing problems at baseline was associated with a mean increase in median time to achieving sitting after treatment of 4.03 months (95% CI: 3.92, 4.15) and 3.13 (95% CI: 3.03, 3.23) months, respectively. Additionally, scoring more than a 0 on the HINE-2 kicking item was associated with a mean decrease in median time to achieving sitting after treatment of 4.14 (95% CI: 4.01, 4.27) months. One of the most pronounced differences in the partial dependence predicted time-to-sitting curves was associated with the ability to roll fully prone from supine, who have a mean difference in median time-to-sitting of 5.76 (95% CI: 5.51, 6.01) months earlier than those who cannot. The full partial dependence for the binary variables is given in Table 2.

**Table 2.** Partial Dependence Summary of Impact on Binary Features on Median Time to Sit.

|  | Median Estimated Time-to-Sit After Treatment<br>(months) |  |
| --- | --- | --- |
| Requiring Intervention? | Yes | No |
| Feeding Support | 19.77 (19.16, 20.29) | 15.65 (14.9, 16.32) |
| Swallowing Difficulties | 20.05 (19.42, 20.59) | 16.88 (16.3, 17.46) |
| Tracheostomy | 18.29 (17.77, 18.83) | 17.77 (17.17, 18.22) |
| Functional Ability? | No | Yes |
| Kick | 19.03 (18.53, 19.57) | 14.8 (14.06, 15.69) |
| Rolling fully Prone from Supine | 18.01 (17.49, 18.46) | 11.66 (9.39, 14.92) |
| Neck Muscle Activation during Pull to Sit | 18.32 (17.89, 18.8) | 14.34 (13.16, 15.77) |
| Antigravity Arm Movement in Side Lying | 18.56 (18.13, 19.1) | 15.84 (15.09, 16.72) |
| Hand Grip when Shoulder Lifted | 18.44 (18.03, 19) | 16.66 (15.98, 17.35) |
| Antigravity Shoulder Movement | 18.57 (18.16, 19.11) | 16.43 (15.78, 17) |
| Lower Limb Movement Proximal to the Ankles | 18.41 (17.8, 19.05) | 17.29 16.69, 17.83) |

Using the model, we also provide the likelihood of sitting for a theoretical patient. We defined functional phenotypes (CHOP-Intend items, HINE-2 kicking, CHOP-INTEND and HINE-2 total scores, and NIV hours) for the patients as ”strong” if they were in the upper third of patients (average 83^rd^ centile), ”intermediate” in the middle third (50^th^ centile), ”weak” if they were between the lower third and bottom sixth (average 25^th^ centile), and ”profoundly weak” if they were in the bottom sixth (average 8.3^rd^ centile). The theoretical patients were attributed the median growth centiles, symptom onset at 4 months and treatment at 6 months. The full patient profiles are presented in Supplementary Table 1. The probability of sitting by time since nusinersen initiation is presented in Figure 5, with the median time to sit in brackets.

**Figure 5.**
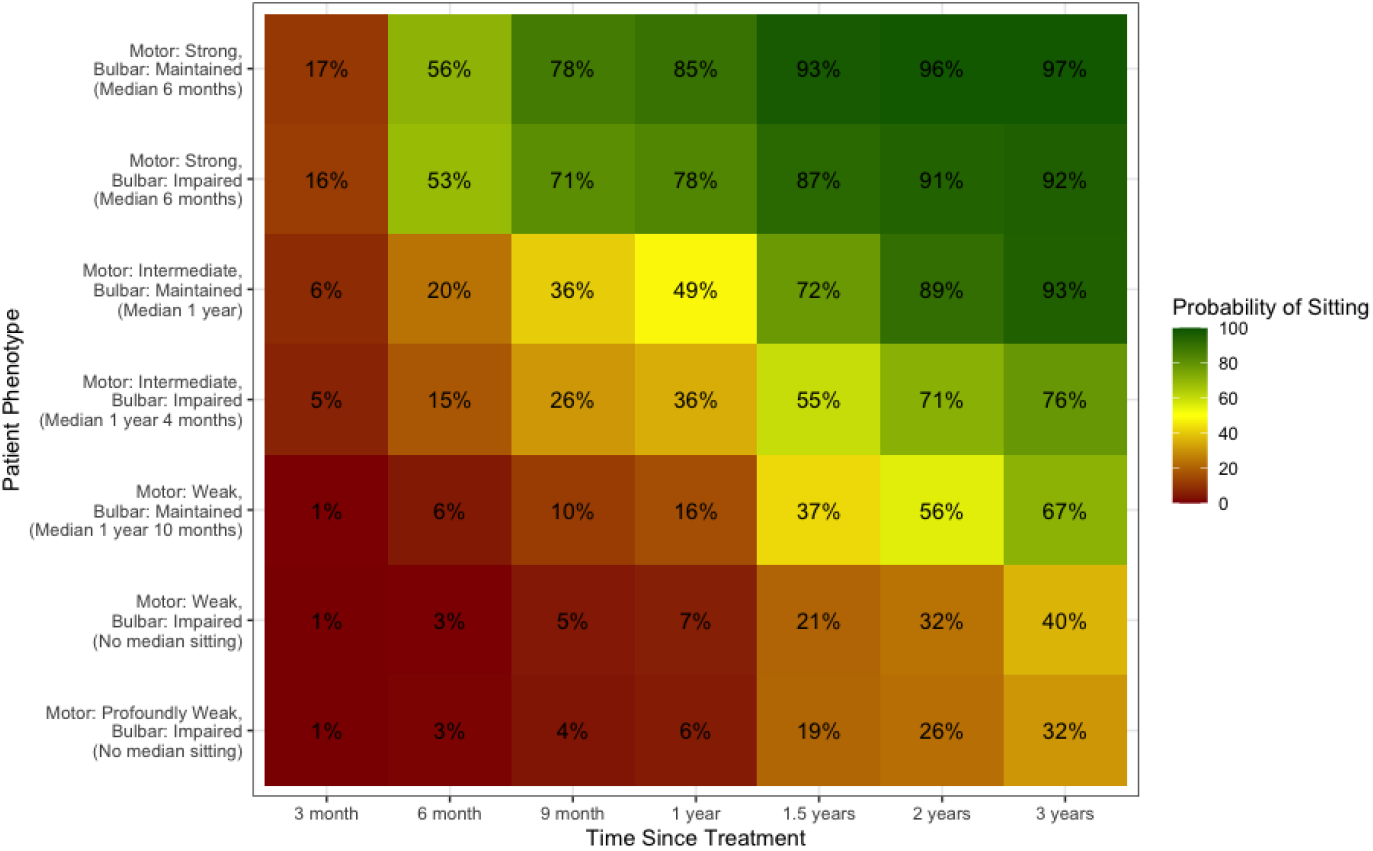
Predicted Likelihood of Sitting by Motor Function and Bulbar Function Presentation

In the strongest patients (those who had already achieved the motor function abilities included in the model), the likelihood of sitting within 6 months of treatment was over 50%, regardless of the patient’s bulbar function, with those with bulbar impairment at baseline displaying only a marginally smaller likelihood across all follow up times. In those with intermediate motor function ability (inability to role fully prone from supine, no antigravity arm movement whilst in side lying and no neck muscle activation in pull to sit, but ability on the other four motor function abilities included), without bulbar impairment, we predicted that 50% would sit within 12 months of treatment, whilst we predicted 50% of patients with this phenotype and bulbar impairment would sit within 16 months of treatment. In weak patients (lower limb movement proximal to the ankles but no ability on the other six abilities included in the model), the impact of bulbar function was much more apparent. In those with no bulbar impairment, the patient had a 50% chance of sitting within 22 months of treatment. However, if the patient had impaired bulbar function, the likelihood of sitting remained low, with only 40% sitting within 4 years of treatment. In the weakest participants who had no ability on any of the motor abilities included in the model, the prognosis was similar, with only 32% of patients sitting within 4 years of treatment.

For patients with intermediate function and bulbar impairment, their inability to kick at baseline (on the HINE-2) was the factor that reduced most the predicted likelihood of sitting over the follow-up period, as can be seen by the large negative values for kicking in Figure 6A. This was particularly true between 1 and 2 years after nusinersen treatment. Additionally, the patient’s swallowing problems at baseline had the largest negative impact on the sitting probability over time. Conversely, in a patient with weak intermediate function but no bulbar impairment, their inability to kick was the factor that reduced most the likelihood of sitting, whilst their maintained bulbar function increased the likelihood of sitting (see Figure 6B). It is worth noting that in these patients, if they were on NIV for at least 10 hours daily, this reduced their likelihood of sitting particularly at 12 months after treatment. Additionally, neck muscle activation in pull-to-sit (CHOP item 14) and ability to roll fully prone from supine (CHOP item 7) had the highest impact locally on the prediction of sitting for the patient with intermediate motor and impaired bulbar function (see Figure 6C), meaning that an identical patient with ability on these items would have a higher predicted likelihood of sitting. Similarly, these items had the largest local impact for the patient with weak motor function and maintained bulbar function (see Figure 6D).

**Figure 6.**
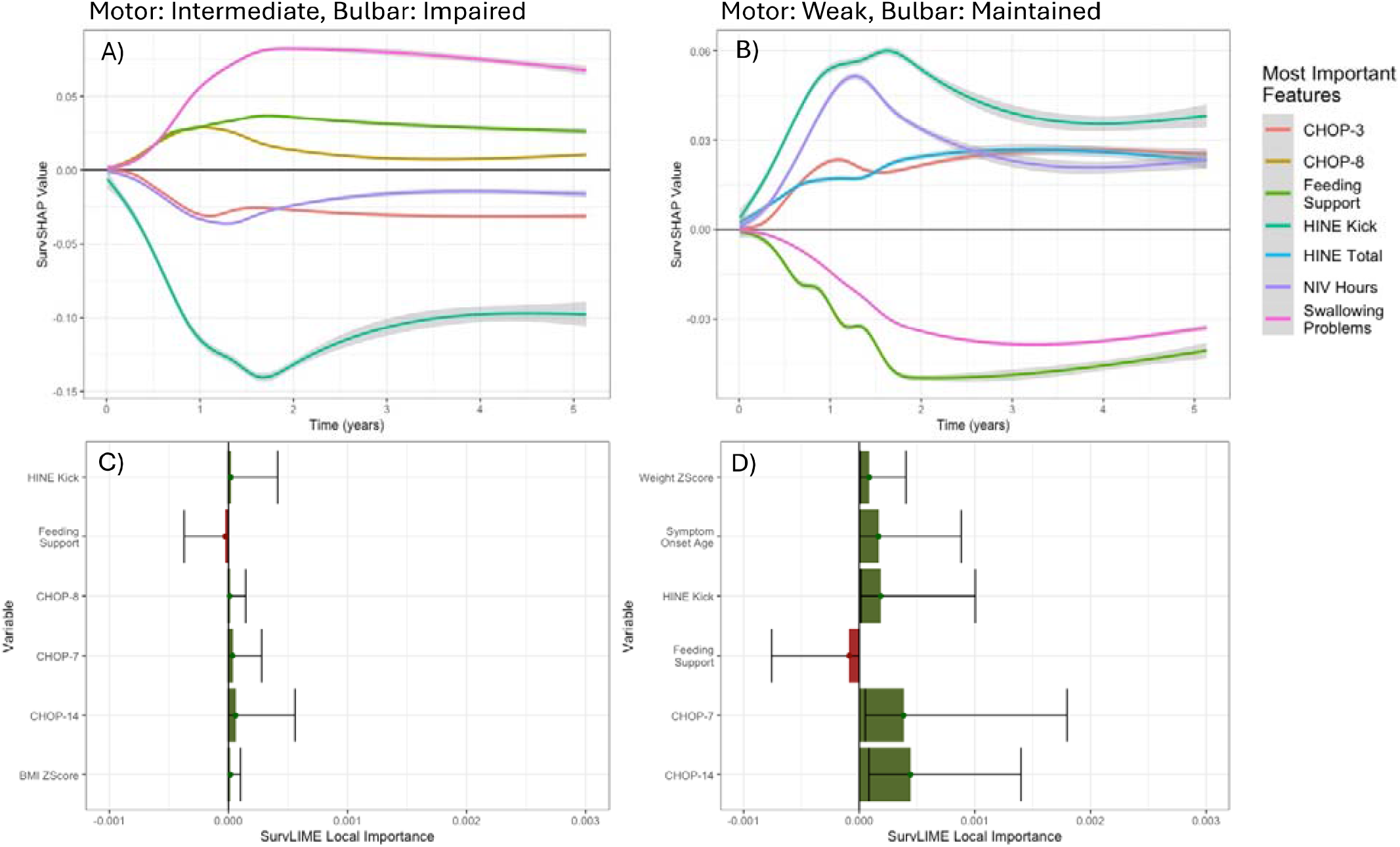
Explainability of Predictions-SurvSHAP and SurvLIME. Note here that the interpretation of SurvSHAP values is converse, in that negative values indicate a higher likelihood of sitting.

## 5 Discussion

This study represents the first attempt at predicting the ability to sit after nusinersen treatment in non-sitters with SMA using machine learning. Machine learning methodologies are uniquely informative over traditional statistical methods, as they induce interactions and non-linearity of the features, and their use is novel in SMA.

Overall, half (50%) of the participants in our cohort were observed as gaining the ability to sit, and the median age of sitting was 2.41 years (IQR: 1.8, NA). We found that bulbar function (feeding support status, and to a lesser extent swallowing problems) had the most influence on the predicted likelihood of sitting. When considered in isolation, the presence of feeding support or swallowing problems at baseline was associated with a 4 and 3-month delay in median time-to-sitting, respectively. Additionally, for predictions of individual patients, the presence of a feeding tube had a high level of influence on the predicted time-to-sitting in those with intermediate and weak baseline function. This is in line with a previous finding, which stated that there was a higher rate of nutritional support required at baseline in those who did not gain the ability to sit within 14 months of treatment ^14^. Rather than feeding support preventing these patients from sitting, we may argue that feeding support status is broadly representative of disease severity at baseline, and indicates that those who are more severely affected at baseline are less likely to sit.

The ability to kick at baseline, assessed as scoring at least 1 on the kicking item of the HINE-2, was also an influential feature in predicting the likelihood of sitting. The total score on the HINE-2 was also included in the model and had some influence, although to a lesser extent. When considered in isolation, scoring at least 1 on the HINE kicking item (i.e. ability to at least kick horizontally) was associated with a median time-to-sit 4 months earlier than in patients who scored 0. A previous finding reported that a baseline score of ≥2 on the HINE-2 was associated with a 300% higher rate of sitting at a 14-month follow-up ^26^. As the ability to kick on the HINE-2 necessarily implies a HINE-2 total score of at least 1, we may hypothesize that the ability on the HINE-2 kicking item was actually the driver of this previously observed result.

Several of the CHOP-INTEND items were found to be predictive of the likelihood of sitting. Primarily, this was antigravity shoulder movement (4 on item 1), neck muscle activation in pull to sitting (at least 2 on item 14), the ability to roll fully prone from supine (at least 3 on item 7), and antigravity arm movement in side-lying (4 on item 8). Most notably, when considered in isolation, patients who were able to roll fully prone from supine had a median predicted time-to-sit after treatment 5.8 months earlier than those who did not. Whilst a difference in total CHOP-INTEND score at baseline has previously been significantly associated with the proportion of patients sitting at 14 months ^26^, the CHOP-INTEND total score had a lower level of influence in our model than the specific items highlighted above.

The requirement of respiratory support as assessed by hours of NIV was also mildly influential in the model, most notably in patients who were weak at baseline. This association is in line with previous findings, which reported a higher rate of NIV use at baseline in those who could not sit at 14 months compared to those who could ^26^. A previous study showed that patients treated between 12 and 24 months who were already on NIV displayed a stabilisation of the CHOP-INTEND trajectory compared to those who did not require NIV support, who showed instead an increase in CHOP-INTEND over time ^27^.

Finally, many of the previous studies on this topic have focused on nusinersen starting age, symptom onset and *SMN2* copy number to explain differential sitting rates ^14,28^. Whilst nusinersen starting age and symptom onset age were selected for inclusion for our model, the inclusion of the *SMN2* copy numbers did not improve the prediction accuracy. The lack of selection of *SMN2* copy numbers may be due to the relative homogeneity of the cohort included in this study, with 74% of patients having 2 *SMN2* copies. Overall, the influence of both symptom onset and nusinersen starting age was small relative to that of feeding support and specific motor function items, highlighting that disease severity at baseline is a much more relevant factor.

This is a preliminary study, and consequently, further development is needed before its findings can be integrated into a clinical decision-making paradigm. In the majority of cases, pre-symptomatic and early onset symptomatic SMA patients have the option to receive any of the three approved SMN enhancing DMT, and as such the replication of this work in those treated with onasemnogene abeparvovec and risdiplam would be crucial to informing decision-making. Additionally, in light of changing patterns of treatment including patients bridging to onasemnogene abeparvovec ^29^, the inclusion of features which capture short-term functional changes would be an important extension to this model.

As with most rare diseases, the number of features relative to the number of patients is very high, and as such this study would benefit from a larger sample size. This study currently includes the majority of non-sitter SMA patients treated with nusinersen in the UK and Italy up to December 2022. Given the increasing heterogeneity in treatment patterns due to the more recent approval of OA and risdiplam, with increasing number of SMA I patients either starting with OA or bridging/switching to OA, it is unlikely that increasing the cutoff date of the sample would increase the sample size. Instead, a wider data collection including other international sites could be helpful in improving the model. To counteract the high number of features compared to the number of participants, we have included multiple featuring ranking methods, including rankings based on correlation, regression and mutual information. However, there may be some features that were excluded from the model due to low feature rankings that would have improved the model fit, but it is computationally infeasible to assess the performance of every permutation of features. Additionally, even though we have employed cross-validation in the training process, the performance of the models in the test data is slightly worse, which may suggest mild overfitting.

A secondary limitation of this model is that it is not possible to understand causality using this methodology. For some features, it is unlikely that there is a causal relationship at play-for example, it is unlikely that a patient’s need for feeding support causes a lower likelihood of sitting, and instead, it is more likely that there is some latent measure of disease severity, which causes both the patient to require feeding support, and to have a lower likelihood of sitting. However, there are other variables where a causative relationship could be possible. In order to model causal relationships between time-to-sitting and baseline covariates, an alternative model structure would be necessary, such as that of a causal structural equation model.

### 4.1 Conclusion

Our study shows that machine learning can predict the patient-specific likelihood of sitting, with an overall good performance. Bulbar function was found to have a significant impact on the predicted likelihood of sitting in all but the strongest patients, with presence of feeding support at baseline associated with a delay in median time to sit of 4 months across the whole cohort. Motor function abilities that are predictive of a higher likelihood of sitting included antigravity shoulder movement, antigravity arm movement whilst in side-lying, rolling fully prone from supine, the ability to kick and activation of neck muscles in pull to sit. These results can inform treatment-response discussions with parents and carers of SMA babies, and serves as the preliminary work for informed treatment decision-making in SMA.

## Data Availability

The anonymised data used in this study can be made available for academic use to interested parties on receipt and approval of an audit request to both SMA Reach UK and the Italian Network.

## Acknowledgements

The author of this publication would like to acknowledge the work of the SMA REACH UK Network for their contribution in the collection and provision of the data. Prof Francesco Muntoni is the Chief Investigator of the SMA REACH UK Project in the UK (REC Reference 13/LO/1748, IRAS project ID: 122521), via UCL and GOSH.

SMA REACH UK would like to acknowledge the contribution of commercial funders Biogen, Roche and Novartis. Historically, funding for the SMA REACH UK Project was provided by the SMA TRUST, and Muscular Dystrophy UK, the MRC Translational Research Centre at UCL and Newcastle (MR/K501074/1) and the National Institute of Health Research Biomedical Research Centre (515048) at Great Ormond Street Hospital for Children NHS Foundation Trust and University College London. The funders had no role in study design, data collection and analysis, decision to publish or preparation of the article.

The support of Famiglie SMA, Telethon (GSP 13002), and ASAMSI to the Nemo Center in Rome and to the Italian network is gratefully acknowledged.

We are grateful to Colm O’Reilly for his work in creating the diagrams that describe the motor function abilities used in the model.

We also acknowledge the Turing-Roche Strategic Partnership, which authors GS, TC and RM are part of.

## 8 Author Contribution

**Georgia Stimpson:** Writing – original draft, Visualization, Methodology, Investigation, Formal analysis, Data curation, Conceptualization, Formal Analysis. **Emer O’Reilly:** Data curation, Conceptualization, Writing – review & editing. **Giorgia Coratti:** Data curation, Writing – review & editing. **Deborah Ridout:** Conceptualization, Writing – review & editing, Supervision, Methodology. **Tapabrata Chakraborti:** Writing – review & editing, Supervision, Methodology. **Robin Mitra:** Writing – review & editing, Supervision, Methodology. **Roberto de Sanctis:** Data curation, Writing – review & editing. **Marika Pane:** Data curation, Writing – review & editing. **Mariacristina Scoto:** Writing – review & editing. **Eugenio Mercuri:** Data curation, Writing – review & editing. **Francesco Muntoni:** Conceptualization, Writing – review & editing, Supervision. **Giovanni Baranello:** Conceptualization, Writing – review & editing, Supervision. The data has been directly accessed by GS and GC.

## 9 Supplemental

Supplementary Document 1: Motor Function Abilities included in the final model

**Supplementary Figure 1.**
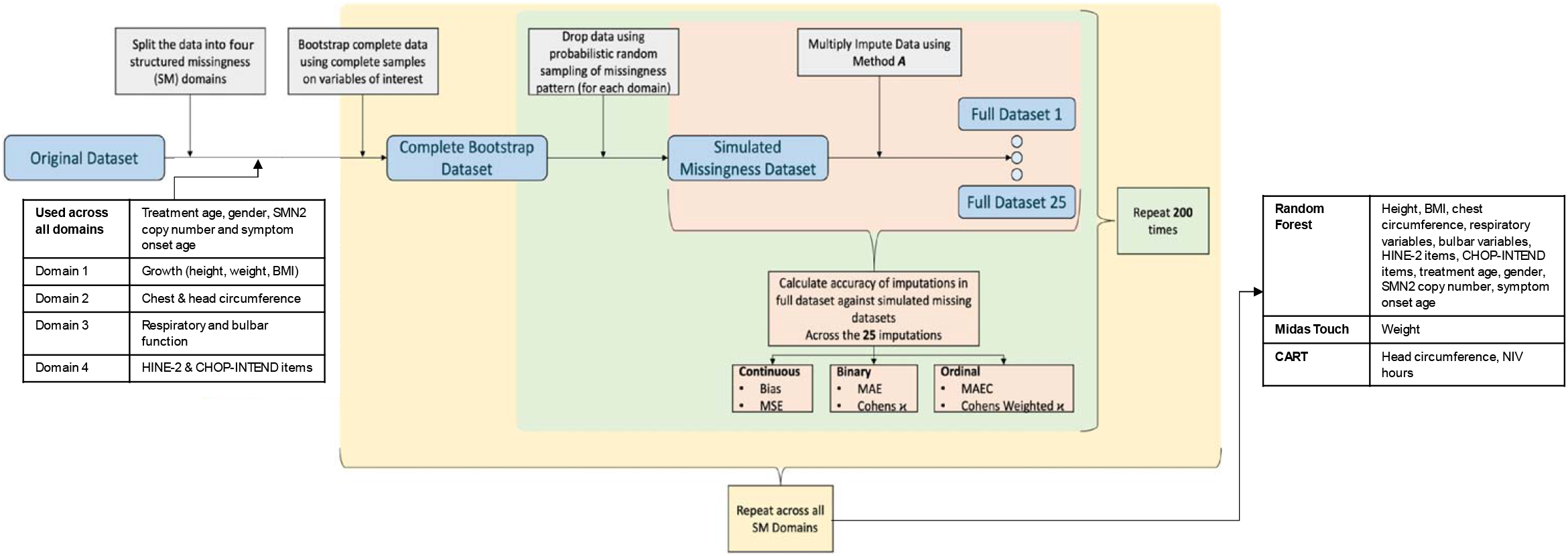
MICE Imputation Selection of Method with respect to Structured Missingness

**Supplementary Table 1.** Patient Profiles for Prediction.

| Motor | Bulbar | Feeding Support | Swallowing Problems | CHOP-2 Score≥2 | HINE-2 Kick Score≥1 | CHOP-3 Score=4 | CHOP-1 Score=4 | CHOP-7 Score≥3 | CHOP-8 Score=4 | CHOP-14 Score≥2 |
| --- | --- | --- | --- | --- | --- | --- | --- | --- | --- | --- |
| Strong | Maintained | No | No | Yes | Yes | Yes | Yes | Yes | Yes | Yes |
| Strong | Impaired | Yes | Yes | Yes | Yes | Yes | Yes | Yes | Yes | Yes |
| Intermediate | Maintained | No | No | Yes | Yes | Yes | Yes | No | No | No |
| Intermediate | Impaired | Yes | Yes | Yes | Yes | Yes | Yes | No | No | No |
| Weak | Maintained | No | No | Yes | No | No | No | No | No | No |
| Weak | Impaired | Yes | Yes | Yes | No | No | No | No | No | No |
| Profoundly Weak | Impaired | Yes | Yes | No | No | No | No | No | No | No |
| Motor | Bulbar | Height Zscore | Weight Zscore | BMI Zscore | HINE-2 Total | CHOP-INTEND Total | NIV Hours | Tracheostomy | Symptom Onset | Treatment Age |
| Strong | Maintained | -0.76 | -1.61 | -1.42 | 3 | 41 | 0 | No | 3 months | 6 months |
| Strong | Impaired | -0.76 | -1.61 | -1.42 | 3 | 41 | 0 | No | 3 months | 6 months |
| Intermediate | Maintained | -0.76 | -1.61 | -1.42 | 1 | 30 | 0 | No | 3 months | 6 months |
| Intermediate | Impaired | -0.76 | -1.61 | -1.42 | 1 | 30 | 0 | No | 3 months | 6 months |
| Weak | Maintained | -0.76 | -1.61 | -1.42 | 0 | 14 | 10 | No | 3 months | 6 months |
| Weak | Impaired | -0.76 | -1.61 | -1.42 | 0 | 14 | 10 | No | 3 months | 6 months |
| Profoundly Weak | Impaired | -0.76 | -1.61 | -1.42 | 0 | 8 | 16 | No | 3 months | 6 months |

**Supplementary Table 2.** Feature Ranking by Filtering Method.

| Variable | Feature Ranking Method |  |  |  |  |
| --- | --- | --- | --- | --- | --- |
|  | Correlation | CARS | MIM | MRMR | DISR |
| Gender | 6 | 40 | 57 | 23 | 40 |
| Nusinersen Start Age | 13 | 20 | 2 | 4 | 2 |
| SMN2 Copy Number | 16 | 72 | 62 | 39 | 56 |
| Symptom Onset Age | 22 | 24 | 9 | 12 | 5 |
| Feeding Assistance | 36 | 3 | 3 | 3 | 6 |
| Swallowing Problems Indicator | 12 | 15 | 12 | 7 | 14 |
| BMI Centile | 39 | 22 | 7 | 8 | 9 |
| Head/Chest Circumference Ratio | 3 | 71 | 60 | 33 | 77 |
| Height Centile | 26 | 70 | 6 | 5 | 8 |
| Weight | 2 | 25 | 8 | 15 | 4 |
| NIV | 72 | 30 | 4 | 2 | 3 |
| NIV Hours | 9 | 9 | 78 | 74 | 75 |
| Salbutamol | 1 | 1 | 79 | 62 | 80 |
| Tracheostomy | 10 | 56 | 36 | 11 | 25 |
| HINE Total | 66 | 17 | 5 | 9 | 7 |
| HINE Head Control -1 | 15 | 48 | 64 | 75 | 67 |
| HINE Head Control -2 | 20 | 50 | 40 | 27 | 34 |
| HINE Kicking -1 | 40 | 13 | 10 | 13 | 12 |
| HINE Kicking-2 | 42 | 33 | 45 | 35 | 44 |
| HINE Rolling-1 | 46 | 65 | 70 | 61 | 71 |
| HINE Rolling-2 | 80 | 36 | 73 | 46 | 73 |
| HINE Rolling-3 | 8 | 35 | 74 | 52 | 76 |
| HINE Sitting-1 | 34 | 6 | 55 | 31 | 50 |
| HINE Sitting-2 | 17 | 76 | 77 | 67 | 78 |
| HINE Voluntary Grasp -1 | 41 | 46 | 53 | 47 | 58 |
| HINE Voluntary Grasp -2 | 48 | 4 | 72 | 64 | 62 |
| HINE Voluntary Grasp -3 | 4 | 11 | 76 | 58 | 72 |
| CHOP-INTEND Total | 65 | 54 | 1 | 1 | 1 |
| CHOP-INTEND Item 1, Score 1 | 7 | 12 | 80 | 73 | 79 |
| CHOP-INTEND Item 1, Score 2 | 49 | 73 | 52 | 63 | 57 |
| CHOP-INTEND Item 1, Score 3 | 74 | 49 | 39 | 43 | 38 |
| CHOP-INTEND Item 1, Score 4 | 37 | 8 | 11 | 16 | 11 |
| CHOP-INTEND Item 2, Score 1 | 43 | 27 | 44 | 28 | 43 |
| CHOP-INTEND Item 2, Score 2 | 33 | 10 | 18 | 17 | 13 |
| CHOP-INTEND Item 2, Score 3 | 31 | 16 | 30 | 40 | 18 |
| CHOP-INTEND Item 2, Score 4 | 76 | 62 | 33 | 20 | 29 |
| CHOP-INTEND Item 3, Score 1 | 45 | 80 | 75 | 79 | 74 |
| CHOP-INTEND Item 3, Score 2 | 73 | 43 | 56 | 71 | 60 |
| CHOP-INTEND Item 3, Score 3 | 70 | 31 | 34 | 42 | 41 |
| CHOP-INTEND Item 3, Score 4 | 19 | 32 | 13 | 10 | 10 |
| CHOP-INTEND Item 4, Score 1 | 47 | 79 | 67 | 69 | 63 |
| CHOP-INTEND Item 4, Score 2 | 61 | 45 | 59 | 76 | 61 |
| CHOP-INTEND Item 4, Score 3 | 62 | 63 | 26 | 49 | 39 |
| CHOP-INTEND Item 4, Score 4 | 30 | 5 | 25 | 41 | 23 |
| CHOP-INTEND Item 5, Score 2 | 64 | 47 | 46 | 57 | 48 |
| CHOP-INTEND Item 5, Score 4 | 14 | 59 | 71 | 78 | 70 |
| CHOP-INTEND Item 6, Score 1 | 58 | 21 | 42 | 66 | 46 |
| CHOP-INTEND Item 6, Score 2 | 68 | 68 | 15 | 24 | 16 |
| CHOP-INTEND Item 6, Score 3 | 78 | 66 | 32 | 22 | 35 |
| CHOP-INTEND Item 6, Score 4 | 79 | 44 | 69 | 59 | 69 |
| CHOP-INTEND Item 7, Score 1 | 69 | 39 | 31 | 56 | 33 |
| CHOP-INTEND Item 7, Score 2 | 28 | 53 | 35 | 48 | 42 |
| CHOP-INTEND Item 7, Score 3 | 52 | 14 | 22 | 6 | 21 |
| CHOP-INTEND Item 7, Score 4 | 5 | 57 | 68 | 55 | 68 |
| CHOP-INTEND Item 8, Score 1 | 59 | 74 | 38 | 72 | 53 |
| CHOP-INTEND Item 8, Score 2 | 71 | 34 | 19 | 38 | 26 |
| CHOP-INTEND Item 8, Score 3 | 50 | 64 | 24 | 60 | 37 |
| CHOP-INTEND Item 8, Score 4 | 38 | 26 | 14 | 14 | 15 |
| CHOP-INTEND Item 9, Score 1 | 75 | 28 | 58 | 80 | 64 |
| CHOP-INTEND Item 9, Score 2 | 32 | 75 | 49 | 77 | 59 |
| CHOP-INTEND Item 9, Score 3 | 57 | 60 | 16 | 32 | 20 |
| CHOP-INTEND Item 9, Score 4 | 25 | 23 | 47 | 37 | 52 |
| CHOP-INTEND Item 10, Score 1 | 35 | 61 | 50 | 45 | 36 |
| CHOP-INTEND Item 10, Score 2 | 44 | 77 | 48 | 70 | 49 |
| CHOP-INTEND Item 10, Score 4 | 18 | 7 | 43 | 26 | 45 |
| CHOP-INTEND Item 11, Score 2 | 56 | 41 | 20 | 19 | 22 |
| CHOP-INTEND Item 11, Score 3 | 27 | 42 | 27 | 36 | 28 |
| CHOP-INTEND Item 11, Score 4 | 67 | 55 | 29 | 34 | 31 |
| CHOP-INTEND Item 12, Score 1 | 51 | 38 | 17 | 30 | 17 |
| CHOP-INTEND Item 12, Score 2 | 77 | 29 | 21 | 51 | 24 |
| CHOP-INTEND Item 12, Score 3 | 54 | 19 | 23 | 21 | 19 |
| CHOP-INTEND Item 12, Score 4 | 29 | 67 | 41 | 29 | 32 |
| CHOP-INTEND Item 13, Score 2 | 23 | 18 | 54 | 68 | 51 |
| CHOP-INTEND Item 13, Score 4 | 21 | 52 | 63 | 53 | 54 |
| CHOP-INTEND Item 14, Score 2 | 24 | 2 | 37 | 18 | 27 |
| CHOP-INTEND Item 14, Score 4 | 53 | 58 | 65 | 50 | 65 |
| CHOP-INTEND Item 15, Score 2 | 60 | 37 | 28 | 25 | 30 |
| CHOP-INTEND Item 15, Score 4 | 63 | 69 | 66 | 65 | 66 |
| CHOP-INTEND Item 16, Score 2 | 11 | 51 | 61 | 54 | 55 |
| CHOP-INTEND Item 16, Score 4 | 55 | 78 | 51 | 44 | 47 |

**Supplementary Table 3.**
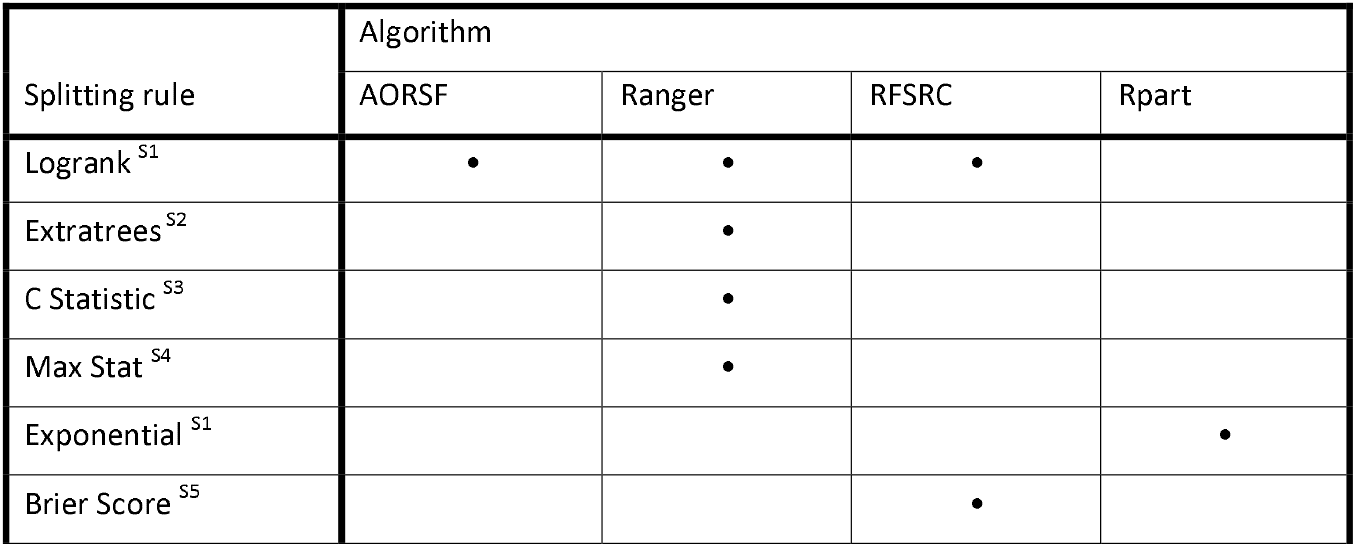
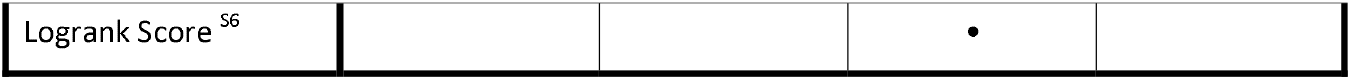
Splitting Rule Implementation by Algorithm.

---

**Supplementary Table 4.** Parameter Name and Domain (I-Integer, R-Real) for Hyperparameter Tuning by Algorithm. Of note, the splitting criterion for AORSF were the critical values of the $\chi_1^2$ test corresponding to significant levels of 0.005-0.1. Additionally, the stopping criteria for the CIT methods reflects a proportion of the total data in the final node, between 2 and 20.

| Algorithm | Hyperparameter Domain |  |  |  |  |
| --- | --- | --- | --- | --- | --- |
|  | Attempted Splits | Splitting Criterion | Stopping Criterion | Candidate Variables | Number of Trees |
| AORSF | nretry<br>I: 0-5 | split_min_stats<br>R: 2.706-7.879 | leaf_min_obs<br>I: 1-20 | mtry<br>I: 1-9 | ntree<br>I: 100-1000 |
| Ranger |  |  | min.bucket<br>I: 1-20 | mtry<br>I: 1-9 | num.trees<br>I: 100-1000 |
| RFSRC |  | nsplit<br>I:1-9 | nodesize<br>I: 1-20 | mtry<br>I: 1-9 | ntree<br>I: 100-1000 |
| Rpart |  | cp<br>R: 0.001-0.1 | minbucket<br>I: 1-20 |  |  |
| CTree | splittry<br>I: 0-5 | Alpha<br>R: 0.01-0.2 | minprob<br>R: 0.021-0.208 |  |  |
| CForest | splittry<br>I: 0-5 | Alpha<br>R: 0.01-0.2 | minprob<br>R: 0.021-0.208 | mtry<br>I: 1-9 | ntree<br>I: 100-1000 |

**Supplementary Figure 2.**
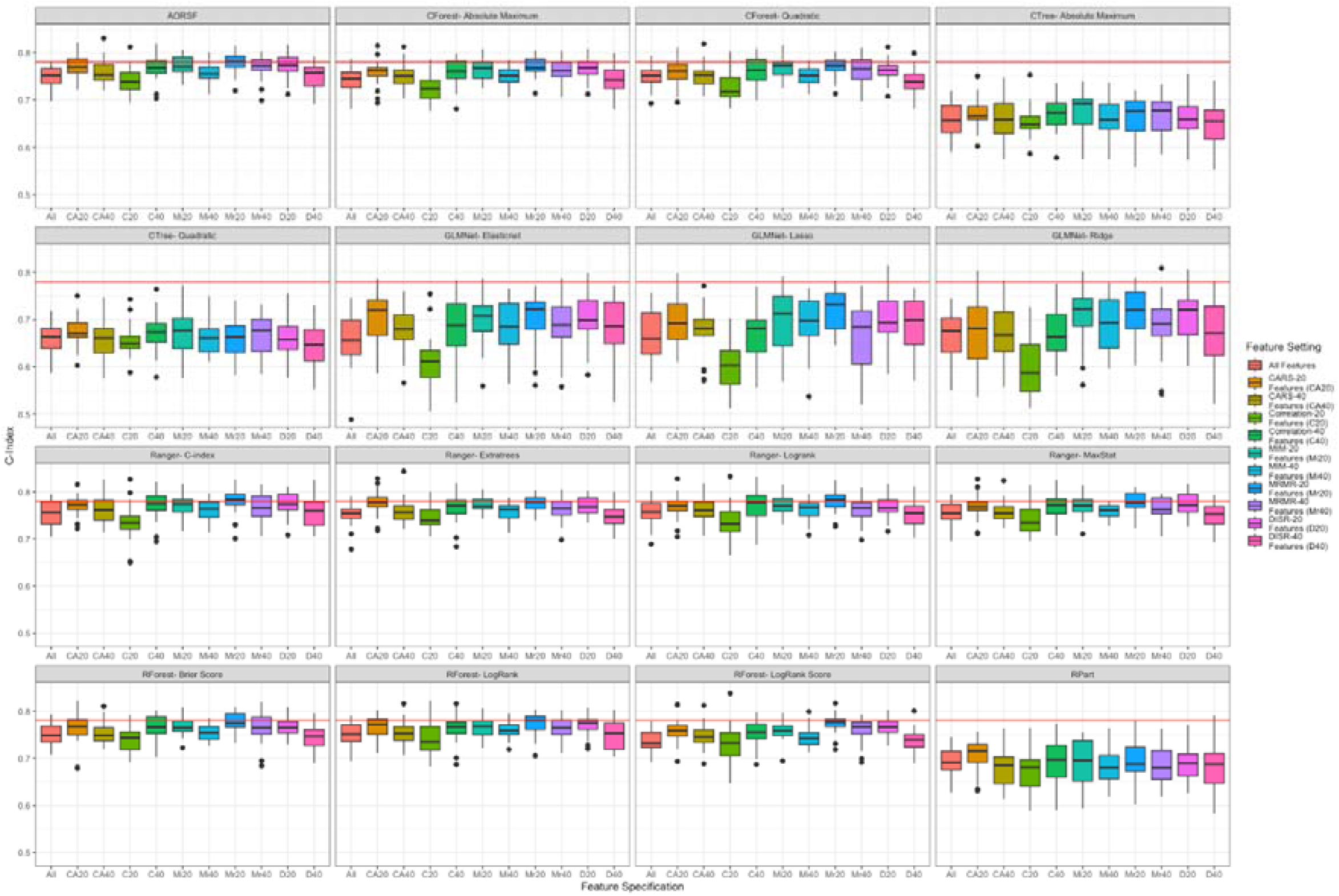
C-Index by Algorithm and Filtering Method

**Supplementary Figure 3.**
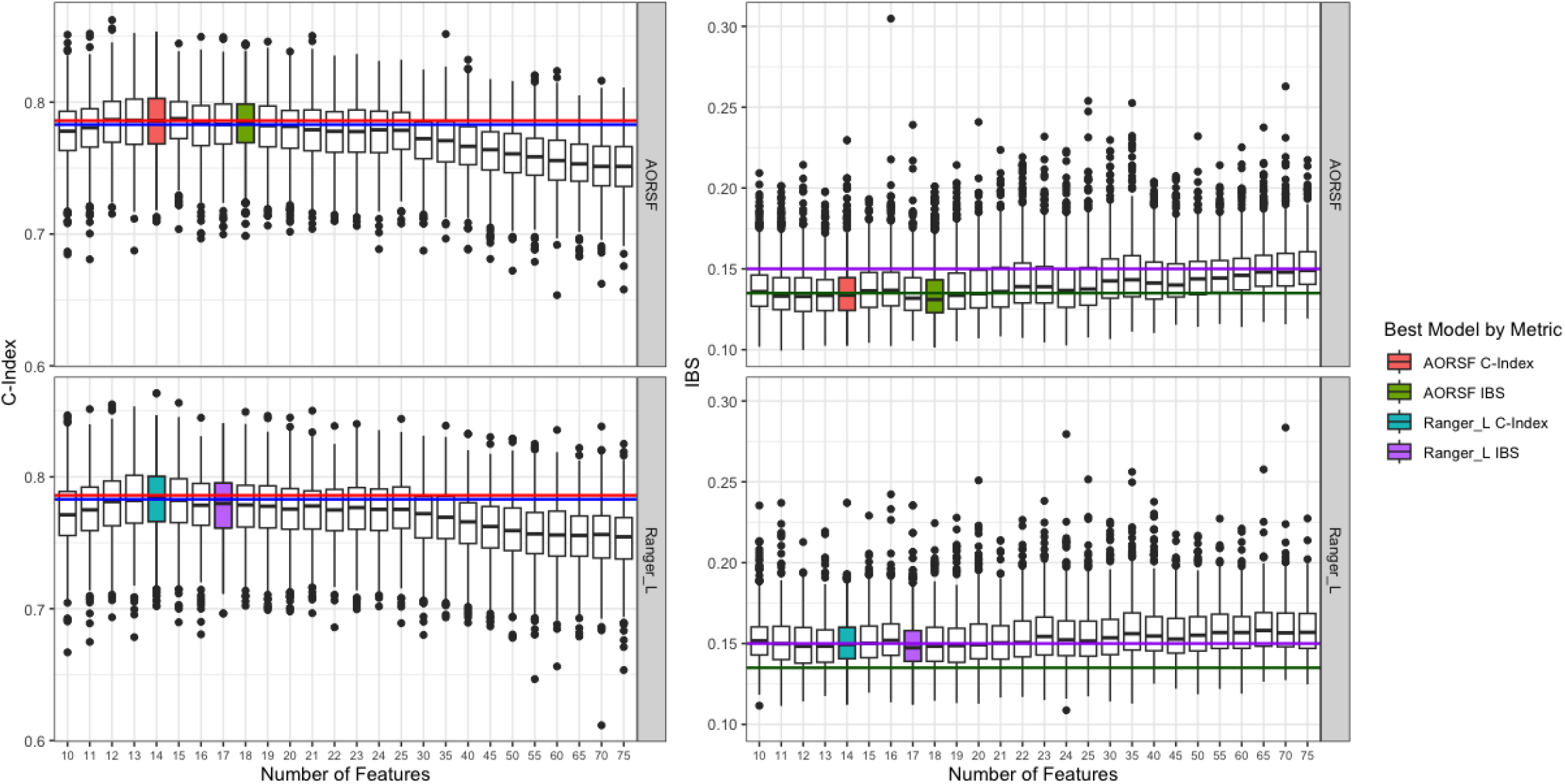
C-Index by Algorithm and Filtering Method for Best Models

